# European lockdowns and the consequences of relaxation during the COVID-19 pandemic

**DOI:** 10.1101/2020.05.19.20106542

**Authors:** David H. Glass

## Abstract

This paper investigates the lockdowns introduced in France, Germany, Italy, Spain, and the UK and also explores the potential consequences of different degrees of relaxation. The analysis employs a two-stage SEIR model with different reproductive numbers pre-and post-lockdown. These parameters are estimated from data on the daily number of confirmed cases in a process that automatically detects the time at which the lockdown became effective. The model is evaluated by considering its predictive accuracy on current data and it is then deployed to explore partial relaxations. The results indicate that the different countries have been successful in reducing the reproductive number to values ranging from 0.67 (95% CI: 0.64 - 0.70) to 0.92 (95% CI: 0.89 - 0.95). Results also suggest that a relaxation of 25% could halt the decline in cases in all five countries, while a 50% relaxation could lead to second peaks that are higher and last longer than the earlier peaks in each country. Even though the relaxations so far may have preserved the success of the lockdowns, vigilance is still needed. A relaxation of around 10-15% is recommended if COVID-19 is to continue to decline in all five countries.

## 1 Introduction

Many countries throughout the world have introduced lockdowns to prevent the rapid spread of COVID-19. How effective have these measures been and how and when should they be relaxed? As many countries have started to relax their lockdowns, this question has become an urgent matter. This paper explores these issues in the context of five European countries: France, Germany, Italy, Spain and the United Kingdom. The approach is to investigate the spread of the virus within these countries both before and after their respective lockdowns took effect and this is achieved by fitting a mathematical model of the spread of infectious disease to data on COVID-19 in each country.

The model in question is a variant of the SEIR model which has been widely used in the modelling of the COVID-19 pandemic [1–8]. This model describes the dynamics of ‘susceptible’ (S), ‘exposed’ (E), ‘infectious’ (I) and ‘recovered’ (R) groups over time. By including the exposed group to model the latent period, it extends the SIR model. SEIR models have been developed in various ways including one that incorporates interactions between different cities in a network [2] and another one that divides the population into different subgroups by age to include differing levels of interaction in society [5].

Here a different approach is adopted since the focus is on using a two-stage SEIR model, with the first stage applying to the period before the lockdown and the second afterwards, and using the available data to learn its parameters. By learning the model from the data on the number of cases, this approach is able to determine the the impact of the lockdowns. The approach assumes that the reproductive number only changes at the lockdown. In this respect, the work is similar to another study of the impact of interventions in European countries that assumed the reproductive number only changed with each intervention, though the approaches differ in other respects [9]. Also, in that study the focus was on the number of deaths rather than the number of confirmed cases which are the primary focus here. However, the results here are compared with corresponding results based on the number of deaths as well as the number of hospital patients in the case of the UK. The model for each country is then evaluated by investigating how well it is able to predict the number of cases of COVID-19 recorded on a given day based on parameters learned from previous days. With the models in place, we are able to compare the effectiveness of the lockdowns in the different countries, make projections for the number of cases in the future and explore the effect of relaxing the restrictions in each country.

## 2 Methodology

Since the goal of the lockdowns is to reduce the transmission rate, *β*, the two-stage SEIR model proposed here involves different values for *β* before and after the lockdown came into effect, but to keep the other parameters fixed. The dynamics of various subgroups of the population before and after a lockdown occurring at *t_lockdown_* are given by the following ordinary differential equations:

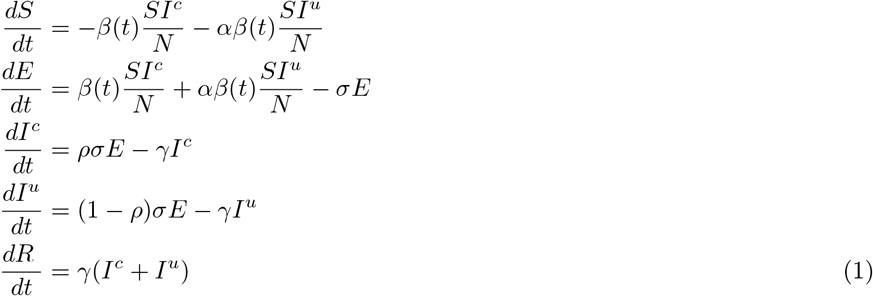

where *β*(*t*) is the transmission rate that has the following values before and after the lockdown

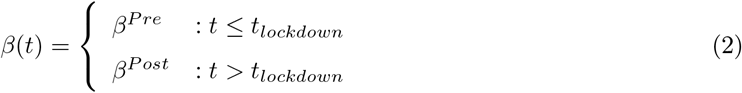

and *N* is the population, *σ* the rate at which those in the exposed group become infectious, *γ* the recovery rate, and *S*, *E*, *I^c^*, *I^u^* and *R* are the susceptible, exposed, infected (confirmed), infected (unconfirmed) and recovered groups respectively, while *ρ* represents the proportion of confirmed cases. In dividing the infected group into two subgroups, we adopt a similar approach to other work that allows one subgroup described as undocumented [2] or sub-clinical / asymptomatic [5] to have a transmission rate reduced by a factor *α*.

The basic reproduction number *R*_0_ is given by *ρβ/γ* + (1 − *ρ*)*αβ/γ*, so alternatively we can say that different values of *R*_0_ are used before and after the lockdown. We can denote these as 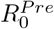, the value before the lockdown, and 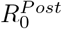, the value afterwards, corresponding to *β^Pre^* and *β^Post^*respectively. The approach is then to learn the values of 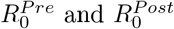 from the daily data on the number of new cases of COVID-19 using the two-stage SEIR model. Data are used from the first day on which there were 100 or more reported new cases confirmed in the country. This date is taken to be day zero and the calculations proceed from there. We also carry out the same process with the data on the number of deaths due to COVID-19 with day zero representing the first day on which there were 10 or more deaths reported in a given country (for details on the data used, see supplementary information). Parameter learning is achieved by integrating the differential equations using the Euler method with a step size of one day and finding parameters that fit the data best in the sense of minimizing the sum of the squared residuals.

We set the parameter *σ* =1 − *e*^−1/3.8^ representing the daily probability of a transition from exposed to infected. This is based on two factors: the incubation period and pre-symptomatic infection. There have been many studies of the incubation period for COVID-19 (see for example [10–12]). A metaanalysis of relevant literature gives a mean incubation period of 5.8 days [13]. There is also evidence of pre-symptomatic transmission of COVID-19 (see for example [14, 15]) with a pre-symptomatic period of infection of about 2 days [16]. Hence, our value for *σ* is based on the difference between these two estimates to give a latent period, *t_l_*, of 3.8 days, which is similar to that used by Li *et al* [2].

The recovery rate, *γ*, is based on the infectious period, *t_i_*, which presents a challenge since a wide range of values have been estimated in the literature (for discussion see [16]). Here, we set *γ* =1−*e*^−1/3.4^ based on estimates of the infectious period in China before and after travel restrictions were introduced of around 3.4 days [2]. This is at the low end of the estimates found in the literature, but it seems justified in the current context for two reasons. First, it is not the infectious period *per se* that is relevant for SEIR models, but the period during which the infection could contribute to transmission. Isolation measures were in place in all the European countries considered here which would have limited the scope for transmission. Second, there is evidence that transmissibility is highest around the onset of symptoms. In a secondary analysis of published data, Casey *et al* [17] suggest that transmission is most likely in the day before symptom onset and estimate that 56.1% of transmission occurs during the pre-symptomatic period based on a pooling of published results (see also [18, 19]). Furthermore, our values for both the latent and infectious periods are in line with estimates of the generation time and serial interval [20]. We also consider other values of several parameters in supplementary material to see how they affect results.

In order to fit the model to the number of newly confirmed cases, we need to identify the proportion (*ρ*) of confirmed cases out of the total number of cases (confirmed and unconfirmed) for a given country. The number of confirmed cases is known and for the total number of cases we divide the number of recorded deaths due to COVID-19 in a given country by a mortality rate of *m* =0.66% [21], though once again we consider other values in supplementary material.

Intuitively, it might seem easy to include the lockdown in a given country. Since the date of day zero is known from the available data and the date of the lockdown is also known, it might be thought that the number of days between day zero and the lockdown could be used to incorporate it in the model. However, there is a delay between the onset of infection and subsequent confirmation. Lin *et al* report a 14 day delay between two datasets with largely the same group of patients [3]. This delay can depend on the availability of testing and on when people seek medical advice, for example, and so can differ from one country to another. Hence, rather than specifying this delay *a priori*, we learn it from the data by determining the value of *t_lockdown_* in equation (2) that gives the best fit (see supplementary material for further details). See table 1 for a summary of the key parameters in the model.

**Table 1:** Summary of key parameters. Results for several other values of *t_l_*, *t_i_* and *m* are explored in supplementary material.

| Parameter | Value |
| --- | --- |
| mean latent period, $t_l$ | 3.8 days |
| mean infectious period, $t_i$ | 3.4 days |
| mortality rate, $m$ | 0.66% |
| transmission reduction factor, $\alpha$ | 1 (see supplementary material) |
| proportion of confirmed cases, $\rho$ | based on $m$ and % deaths in given country |
| pre-lockdown reproductive number, $R_0^{Pre}$ | estimated from fitting model to data |
| pre-lockdown reproductive number, $R_0^{Post}$ | estimated from fitting model to data |
| initial number of exposed cases, $E_0$ | estimated from fitting model to data |
| initial number of infections, $I_0^c$ and $I_0^u$ | sum equal to $E_0$ and proportions based on $\rho$ |
| effective date of lockdown, $t_{lockdown}$ | estimated from fitting model to data |

In addition to fitting the two-stage model to the data, we also evaluate it by determining its predictive accuracy. This is achieved using time series cross-validation [22] and lets us see how well the two-stage model generalizes to unseen data, which is relevant for the final part of the paper where we explore the consequences of relaxing the lockdowns. In effect, this amounts to extending the two-stage model to a three-stage version. The two-stage component is used first to learn the pre-and post-lockdown parameters (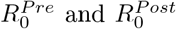) and the value of *t_lockdown_* automatically from the data. After that, the simulation runs until the specified time of the relaxation where 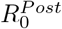 is the pre-relaxation value of and a new post-relaxation value is introduced.

## 3 Results

This section presents results for each of the five countries. Information about day zero, the date the lockdowns were introduced and the proportion of deaths in each country is presented in table 2.

**Table 2:** Dates for day zero (the first day on which at least 100 confirmed new cases were reported), the lockdowns (see [23] and references therein), and the number of deaths as a percentage of the total number of confirmed cases.

|  | Day zero | Lockdown | % deaths |
| --- | --- | --- | --- |
| France | 05/03/20 | 17/03/20 | 19.0 |
| Germany | 05/03/20 | 22/03/20 | 4.7 |
| Italy | 27/02/20 | 09/03/20 | 14.4 |
| Spain | 05/03/20 | 14/03/20 | 11.6 |
| UK | 12/03/20 | 23/03/20 | 14.1 |

### 3.1 Fitting the two-stage SEIR model to the data

Figure 1 presents the two-stage SEIR model that gives the best fit to the daily number of confirmed cases following day zero for each country. In all cases, the model fits the data reasonably well and this is quantified by the *R*^2^ values which highlight that the model fits the Italian data best, whereas the fit for France is poorest. For each country, the impact of lockdown is evident and the two-stage model captures the resulting effect on the number of confirmed cases.

**Figure 1:**
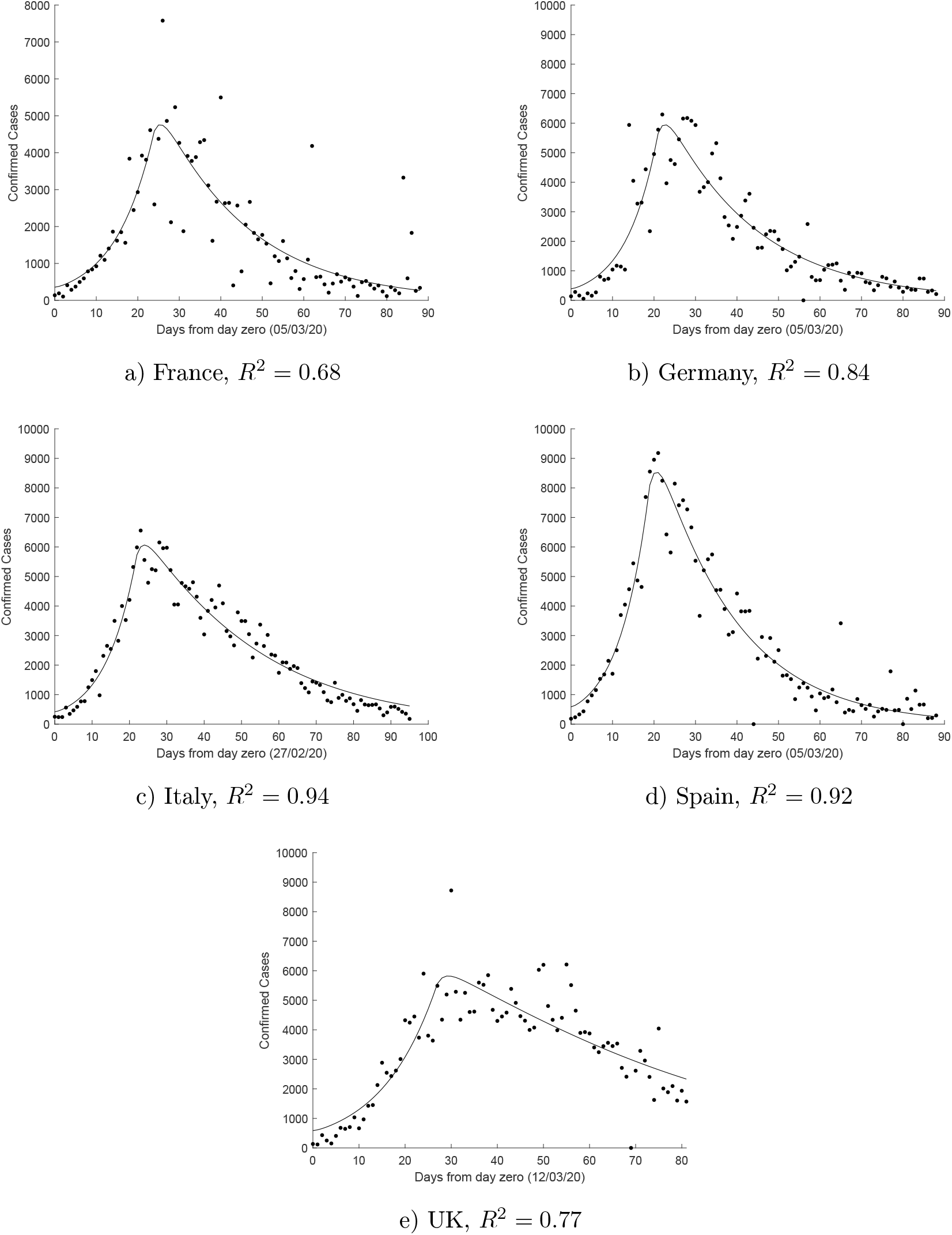
Two-stage SEIR models fitted to the daily confirmed cases for each of the five countries.

It should be noted that it is not the goal of this work to estimate *R*_0_ at the earliest stages of the outbreak of the pandemic in each of the countries. In fact, from figure 1 it can be seen that the model slightly overestimates the number of cases at the earliest stage (before day ten). Hence, the prelockdown *R*_0_ values, 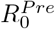, represent situation for about two weeks prior to the lockdown in each country. Nevertheless, it is interesting to note how well the model fits the data over the whole time period shown for each country, suggesting that the assumption of a single *R*_0_ number pre-lockdown and another one post-lockdown is not unreasonable. In particular, the model continues to fit the data well as we move well beyond the lockdown. Interestingly, despite the fact that all of these countries have started to ease their respective lockdowns since early May, 2020, there is no evidence so far from the number of cases of an increase in *R*_0_.

The pre-and post-lockdown reproductive numbers as given by 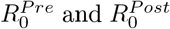 are presented in table 3, together with *R*^2^ values (and RMSE and MAE which will be important later). The pre-lockdown values in table 3 are consistent with other results found in the literature [2, 4, 7, 11]. The results are somewhat lower than those found in an earlier study of European countries by Flaxman *et al* [9], though their results were for initial values whereas ours are for the pre-lockdown period. Furthermore, it also needs to be noted that the results for the 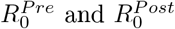 values depend to some extent on the choice of parameters used. We explore this issue in supplementary material. The earlier study found initial evidence that the interventions had a substantial impact on transmission, though at that stage it was too early to say just how effective they had been [9]. The picture has now become clearer with all the reproductive numbers below one. In supplementary material, we also present corresponding results for 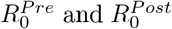 values obtained by fitting the two-stage model to the number of recorded deaths. While there are some differences, particularly where the *R*^2^ value is low, the results in table 3 are similar to those based on the number of deaths, especially for 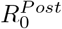. This confirms the general picture that all the post-lockdown values are not only lower, but less than one. Hence, in that sense all the lockdowns have been successful, though at 0.92 the estimate for the UK value of 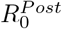 is higher than would have been hoped. According to these results, Spain’s lockdown was the most successful in terms of reducing the reproductive number by the greatest amount, though overall Germany succeeded in keeping the number of deaths much lower than other countries (see figure S1 in supplementary material).

**Table 3:** Estimates for the pre-and post-lockdown reproduction numbers with 95% confidence intervals.

| | $R_0^{Pre}$ | $R_0^{Post}$ | $R^2$ | RMSE | MAE |
| --- | --- | --- | --- | --- | --- |
| France | 2.21 (1.79-2.62) | 0.69 (0.63-0.76) | 0.68 | 891 | 503 |
| Germany | 2.50 (2.10-2.89) | 0.68 (0.63-0.72) | 0.84 | 753 | 496 |
| Italy | 2.39 (2.19-2.59) | 0.81 (0.79-0.82) | 0.94 | 442 | 359 |
| Spain | 2.70 (2.40-3.00) | 0.67 (0.64-0.70) | 0.92 | 707 | 482 |
| UK | 1.90 (1.68-2.11) | 0.92 (0.89-0.95) | 0.77 | 862 | 629 |

Related to the higher 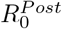 value for the UK, note that the peak in figure 1 for the UK is less pronounced than it is for the other countries. Also, fitting the model to the number of deaths gives a lower value of 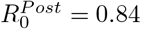 for the UK. To explore the situation in the UK further, an application of the model to UK hospital data is considered in supplementary material. These results are consistent with a higher value for 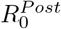 of around 0.9.

### 3.2 Predictive accuracy

Assessing predictive accuracy is relevant here since we will use the two-stage model in section 4 to explore the consequences of relaxing the lockdowns. As noted earlier, the approach adopted is that of time series cross-validation. In each case the last 10 data points are used for testing and *k*-step ahead prediction is used. That is, the model is learned from data up to *k* days before the day that is to be predicted. The two-stage model is evaluated using the metrics RMSE and MAE (see supplementary material for definitions).

Table 4 presents results for *k*-step prediction with values of *k* =5 and 10. It is instructive to compare the values of these metrics for the two-stage model in table 4 with those for all the data for a given country (see table 3). If the results are much poorer on the former than on the latter, that could highlight a potential concern with overfitting and hence for using the model for prediction. Note that the RMSE and MAE values for prediction in table 4 are lower than those in table 3 for France, Germany, Italy and Spain, which is encouraging. While the prediction results for the UK are slightly higher compared to those in table 3, they are not too dissimilar and the higher values can be explained (see supplementary material). The main focus here is not simply on maximizing predictive accuracy since it may well be possible to do that by ignoring the pre-lockdown phase altogether and just fitting models to the post-lockdown data. Instead, the goal is to evaluate the two-stage SEIR model, which can then provide a basis for exploring the consequences of relaxing the lockdowns. The results presented so far give us confidence that it captures the lockdown transition and can be used to make reasonable predictions.

**Table 4:**
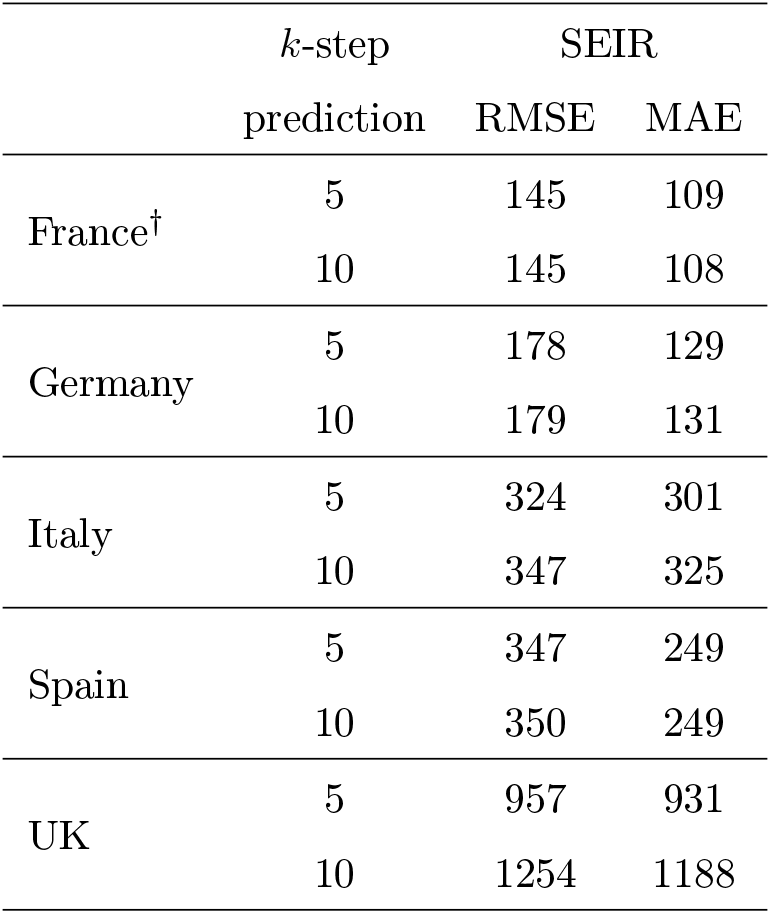
Results for *k*-step ahead prediction for the number of confirmed cases on the last 10 days. † These results exclude two outliers.

| | $k$-step | SEIR | |
| --- | --- | --- | --- |
|  | prediction | RMSE | MAE |
| France<sup>†</sup> | 5 | 145 | 109 |
|  | 10 | 145 | 108 |
| Germany | 5 | 178 | 129 |
|  | 10 | 179 | 131 |
| Italy | 5 | 324 | 301 |
|  | 10 | 347 | 325 |
| Spain | 5 | 347 | 249 |
|  | 10 | 350 | 249 |
| UK | 5 | 957 | 931 |
|  | 10 | 1254 | 1188 |

## 4 Effect of relaxing the lockdowns

Having evaluated the two-stage SEIR model, we now use it to explore the potential effects of partially relaxing the lockdowns in the different countries. Since early May, 2020, some easing of the restrictions has already taken place in all of the countries and and based on the data considered earlier, this does not seem to have affected infection rates to a significant extent. Here we assume that that will continue to be the case until more significant relaxations are introduced towards the end of June, 2020. The different levels of relaxation are implemented on the assumption that they would take effect in terms of daily confirmed cases on 30th June in each of the countries. Based on our results for the time delay between onset of infection and confirmation (see supplementary material), this corresponds to the introduction of partial relaxation of the lockdowns from about 20th June.

Just as the lockdowns were represented as a change in the reproductive number at a single point in time, the same assumption is made for relaxing the lockdowns. The idea is to model relaxations by increasing the reproductive number by a percentage of the difference between the pre-and post-lockdown values. This translates into a corresponding change in the average number of interactions in society, so it is straightforward to interpret. No doubt, there are limitations to this approach since, for example, it does not take into account different measures that could be put in place for different groups within society. However, these limitations are also relevant to the lockdowns in the first place and yet the model represents the lockdowns quite well. Three scenarios are considered:

i. no relaxation - keeping the lockdown fully in place, which is modelled by leaving the *R*_0_ value unchanged at its post-lockdown value, 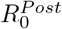,
ii. 25% relaxation - relaxing the lockdown by one quarter so that *R*_0_ increases by 25% of the way back up to the pre-lockdown value, i.e. to a value 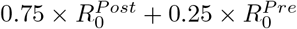, and
iii. 50% relaxation - relaxing the lockdown by one half so that *R*_0_ increases by 50% of the way back up to the pre-lockdown value, i.e. to a value 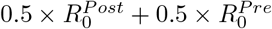.

Needless to say, a lot of caution is required with such attempts to model the future course of COVID-19. Many assumptions have been made about the model. For example, the values of several parameters and the use of a two-stage SEIR model with a single reproductive number before the lockdown and another one afterwards. Also, given the simplicity of the two-stage SEIR model, it cannot be used to model specific relaxations such as allowing schools to re-open. Hence, this approach would need to be complemented by other work that provides more detailed models of society, which could be used to identify what changes would lead to relaxations of a specified percentage and how this might relate to vulnerable groups within society. Nevertheless, the simplicity of the model can also have an advantage becauseitrequiresrelativelyfewmodelassumptionsandyetcanmodelthelockdownsandhenceplausibly relaxations of them too.

Results are shown for numbers of daily confirmed cases and include estimates for two months from the end of June. Not surprisingly given the assumptions, the number of cases continues to fall if there is no relaxation. By the end of June, these numbers are low (below about 200) except for the UK where they are around 1200 cases per day. Although not immediately apparent given the low numbers involved, a 25% relaxation brings to an end the decline in numbers in all five countries, with numbers remaining approximately the same until the end of August in France and rising for the same period in the other countries. The difference between a 25% and 50% relaxation is dramatic with the latter leading to a significant increase in the number of cases in all five countries by the end of August. The estimates suggest that the numbers could be back to around half of what they were at the time of the first peak in Germany and Italy and close to the value at the first peak in the UK.

Sensitivity analysis of these results involved considering different values for the parameters for the latent period, infectious period and mortality rate and results are presented in supplementary material. These results confirm the general picture presented here. For the various parameters considered, increasing the mortality rate (to 1.32%) yields the greatest increase in the number of cases as a result of relaxation. Hence, results based on this value are compared with the original parameter settings in more detail in figure 3, where we let the models run for 600 days from day zero to get an idea what the longer term consequences might be if there were no further interventions. Of course, a lot of caution is needed with these results, but the general pattern is clear. For each country, 50% relaxation leads to a second peak that is higher and lasts for longer than the first peak for both values of the mortality rate, *m*. Typically, the peak is about twice as high when the mortality rate is doubled from 0.66% to 1.32%. Needless to say, lockdowns would be reintroduced before any of these extremely serious scenarios could occur, but the results highlight that numbers could increase very quickly in some cases. The extremely high result for Germany arises from the assumption that fewer people have had COVID-19 so far in Germany based on the much lower number of deaths and so the effective reproductive number remains higher for longer in Germany.

Finally, supplementary material also investigates the application of the two-stage model to UK numbers of COVID-19 hospital patients. The goodness of fit is very high at *R*^2^ =0.987 and these results should also be more reliable since they are not affected by the level of testing. Worryingly, they suggest that the results in figure 2 may be an underestimate, at least in the UK, since by the end of August the numbers are almost twice what they were at the time of the first peak for a 50% relaxation. The results also suggest that a relaxation of 10-15% would be needed to enable the numbers to continue to decline (see supplementary material).

**Figure 2:**
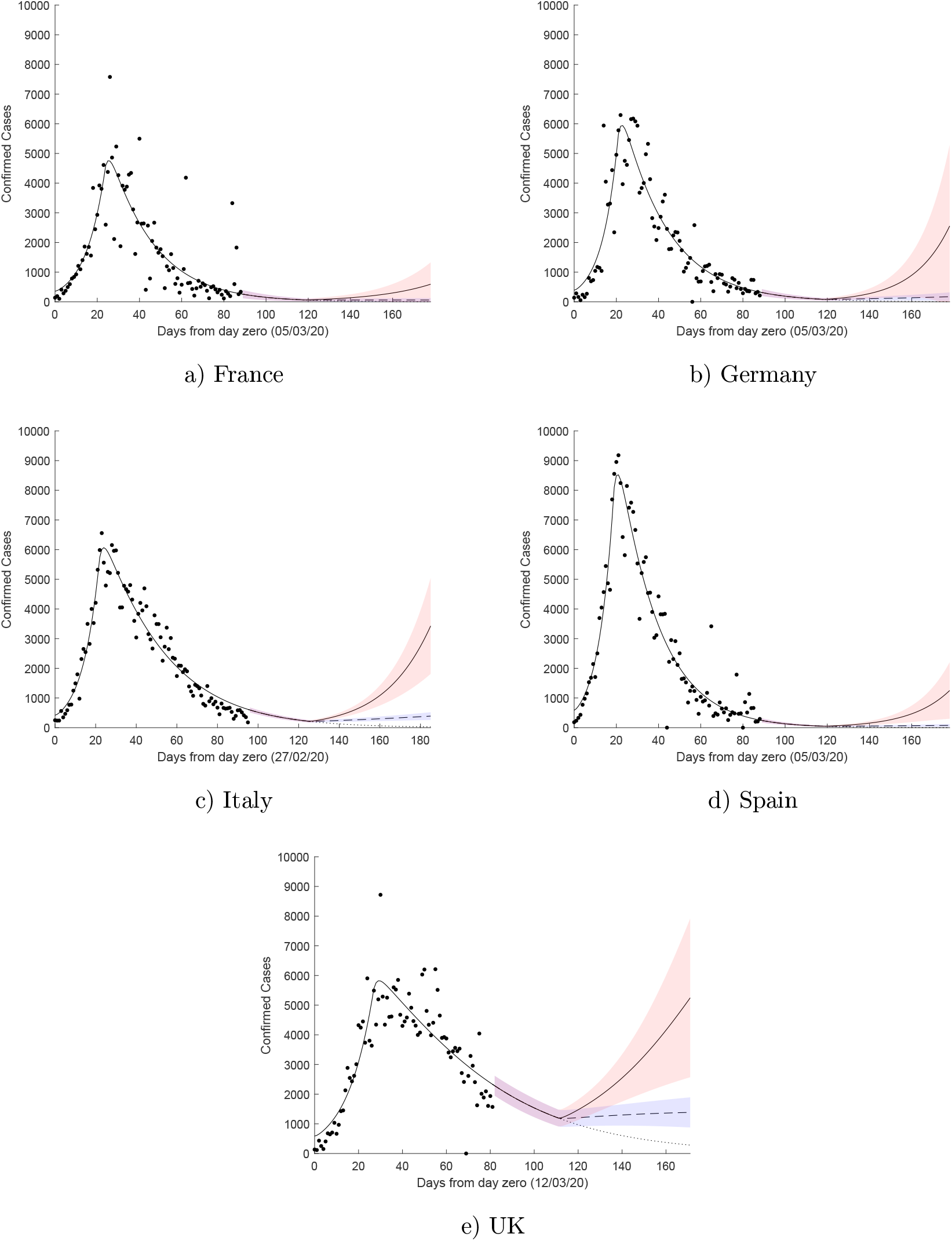
The effect on the daily number of confirmed cases (hospital numbers for the UK) of keeping the lockdown fully in place (·····), relaxing it by 25% (---) and relaxing it by 50% (––). Shaded regions represent 95% confidence intervals. In each case the relaxation is assumed to take effect by 30/06/20, which corresponds to the implementation of the relaxation around 20/06/20.

**Figure 3:**
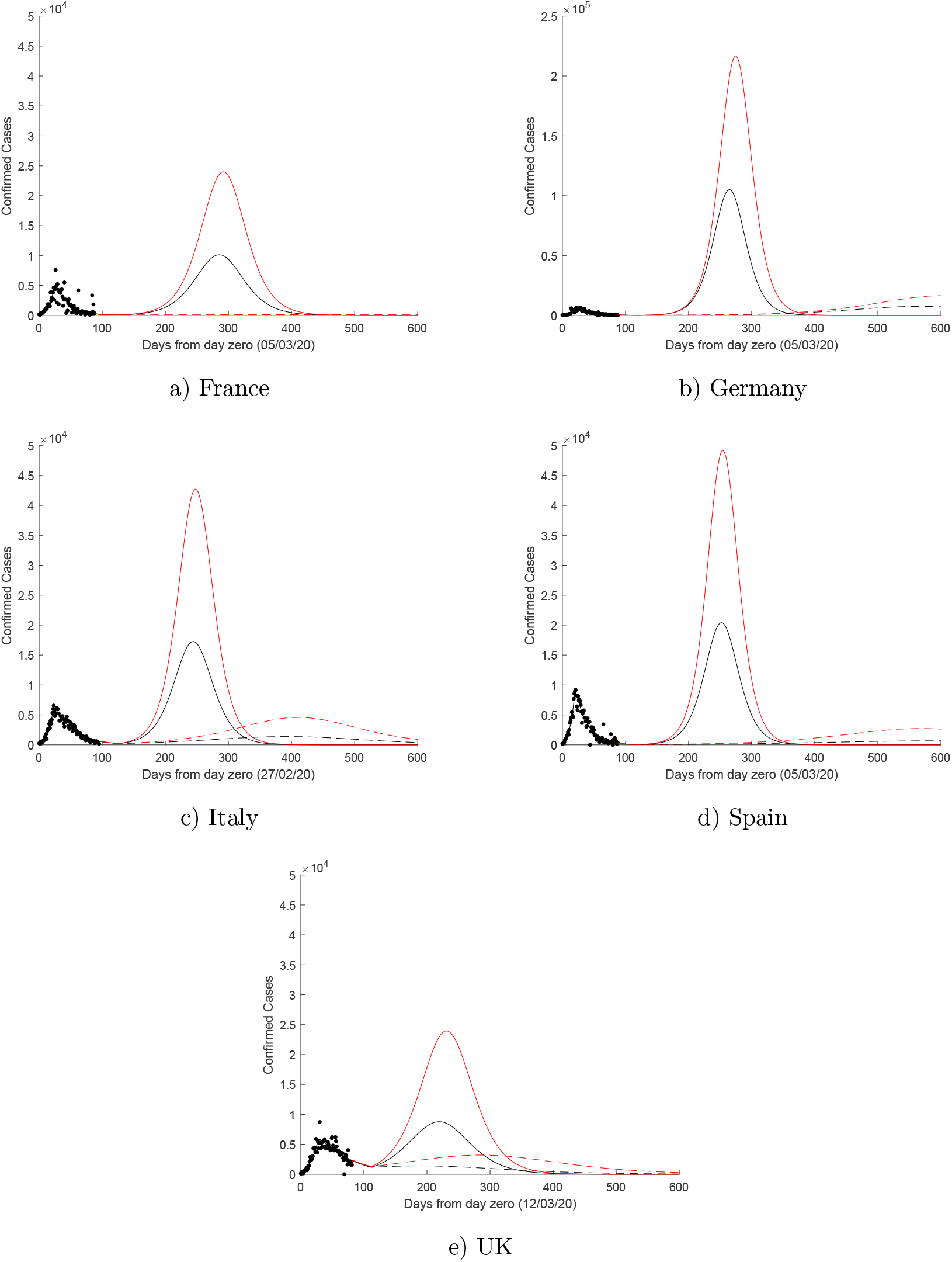
The effect on the daily number of confirmed cases of relaxing the lockdowns assuming a mortality rate of *m* = 0.66% for 25% (- - -) and 50% (––) relaxations, or with *m* = 1.32% for 25% (- - -) and 50% (––) relaxations. Other parameters are as described in table 1. Note that the results for Germany are on a different scale from the other countries.

## 5 Conclusion

A two-stage SEIR model has been fitted to data on the daily numbers of confirmed cases of COVID-19 in France, Germany, Italy, Spain and the UK. This approach enables the date at which the lockdown became effective and the reproduction number before and after the lockdown to be estimated from the data. This enables the effectiveness of the lockdowns to be evaluated. According to these estimates, Spain saw the greatest reduction in the reproductive number.

As well as fitting the two-stage SEIR model to the data, its predictive performance was evaluated using time series cross-validation. Based on this evaluation, the model was used to investigate potential consequences of relaxing the lockdowns towards the end of June. The results indicate that there is a substantial difference between 25% and 50% relaxations. The results suggest that the latter would be disastrous and could lead to a second peak that is higher and lasts longer than the earlier peak in each country, if no further measures were put in place. While the former would not be so serious, it would halt the decline in numbers. A relaxation of around 10-15%, which translates into the level of interactions within society, would be recommended if numbers are to continue to decline in all five countries. Of course, this only applies to society as a whole and particular measures would need to be put in place to protect more vulnerable groups.

Since the two-stage model has been shown to model the lockdowns reasonably well, it can also be used to model relaxations. Hence, as data starts to become available as a result of relaxations being introduced it should be possible to analyze them using the model. Inevitably various assumptions have to be made in the modelling and while the two-stage SEIR model has merits, it is also limited in certain respects. As such, this work could be extended in various ways and used to complement models that take into account age-stratification, for example [5, 24]. Nevertheless, the two-stage model has been evaluated here using the COVID-19 data available so far in five European countries. Hence, caution is needed with any relaxation of lockdowns on the timescales considered in this paper.

## Data Availability

All data used are publicly available.

